# A Deployable Explainable Deep Learning System for Tuberculosis Detection from Chest X-Rays in Resource-Constrained High-Burden Settings

**DOI:** 10.64898/2026.03.31.26349662

**Authors:** John Agumba, Soita Erick, Anthony Pembere, John Nyongesa

## Abstract

**Objectives:** To develop and evaluate a deployable deep learning system with Gradient-weighted Class Activation Mapping (Grad-CAM) for tuberculosis screening from chest radiographs and to assess its classification performance and explainability across desktop and mobile deployment platforms.

**Materials and methods:** This study used publicly available chest X-ray datasets containing Normal and Tuberculosis images. A DenseNet121-based transfer learning model was trained using stratified training, validation, and test splits with data augmentation and class weighting. Model performance was evaluated using accuracy, precision, recall, F1 score, receiver operating characteristic (ROC) curve, and area under the ROC curve (AUC). Grad-CAM was used to visualize regions influencing model predictions. The trained model was converted to TensorFlow Lite and deployed in both a Windows desktop application and a Flutter-based mobile application for offline inference and visualization.

**Results:** The model demonstrated strong classification performance on the independent test dataset, with high accuracy and AUC values indicating effective discrimination between Normal and Tuberculosis cases. Grad-CAM visualizations showed that the model focused primarily on anatomically relevant lung regions, particularly the upper and mid-lung fields in Tuberculosis cases. Deployment testing confirmed consistent prediction outputs and Grad-CAM visualizations across both Windows and mobile platforms.

**Conclusion:** The proposed deployable deep learning system with Grad-CAM provides accurate and interpretable tuberculosis screening from chest radiographs and demonstrates feasibility for offline mobile and desktop deployment. This approach has potential as an artificial intelligence-assisted screening and decision support tool in radiology, particularly in resource-limited and remote healthcare settings.

## Introduction

Tuberculosis is one of the leading infectious causes of death worldwide and continues to place a heavy burden on health systems, particularly in low- and middle-income countries. Early screening and timely diagnosis are essential for reducing transmission, initiating treatment, and improving outcomes. However, access to rapid diagnostic infrastructure remains uneven, especially in rural and under-resourced settings where laboratory testing, radiological expertise, and specialist referral pathways may be limited.

Chest radiography is widely used in tuberculosis screening because it is fast, non-invasive, and routinely available in many clinical environments. Nevertheless, chest X-ray interpretation is dependent on trained readers, and diagnostic consistency may vary across observers and clinical settings. These limitations have motivated the development of artificial intelligence systems capable of assisting healthcare workers by automatically identifying suspicious radiographic patterns and prioritizing high-risk cases for review.

Deep learning, particularly convolutional neural networks, has demonstrated strong performance in medical image analysis. In chest radiography, deep learning models have shown promise in identifying pulmonary tuberculosis and other thoracic abnormalities from image data (1,2). Transfer learning has become particularly important in this domain because medical imaging datasets are often smaller than general computer vision datasets, making initialization from large pre-trained models advantageous (3). DenseNet architectures are especially effective because dense connectivity improves gradient propagation and feature reuse while reducing redundancy in learned representations (4).

Despite these advances, many artificial intelligence solutions for tuberculosis classification remain confined to experimental settings. In addition, most deep learning systems provide only a classification result, without indicating which image regions drove the decision. This lack of transparency limits trust, interpretability, and potential clinical adoption. Explainable artificial intelligence methods such as Grad-CAM provide a practical approach to addressing this limitation by highlighting regions of the image that most strongly contributed to the model output (5).

Another major barrier is deployment. Many published models depend on cloud-based inference or high-performance computing resources and are therefore poorly suited for remote or bandwidth-limited settings. Edge deployment on mobile and desktop devices is important for real-world screening systems because it enables offline use and broad accessibility. TensorFlow Lite offers a pathway for lightweight inference on such platforms (6).

This study presents TBAI Africa, a deployable and explainable deep learning system for tuberculosis screening from chest radiographs. The study aimed to develop a DenseNet121-based binary classification model, integrate Grad-CAM visual explanation, and deploy the resulting system to both mobile and Windows desktop platforms for offline decision support.

## Materials and methods

### Study design

The study followed an end-to-end artificial intelligence workflow consisting of dataset preparation, preprocessing, transfer learning model development, optimization, evaluation, explainable visualization, TensorFlow Lite conversion, and deployment as shown in Figure 1.

**Figure 1:**
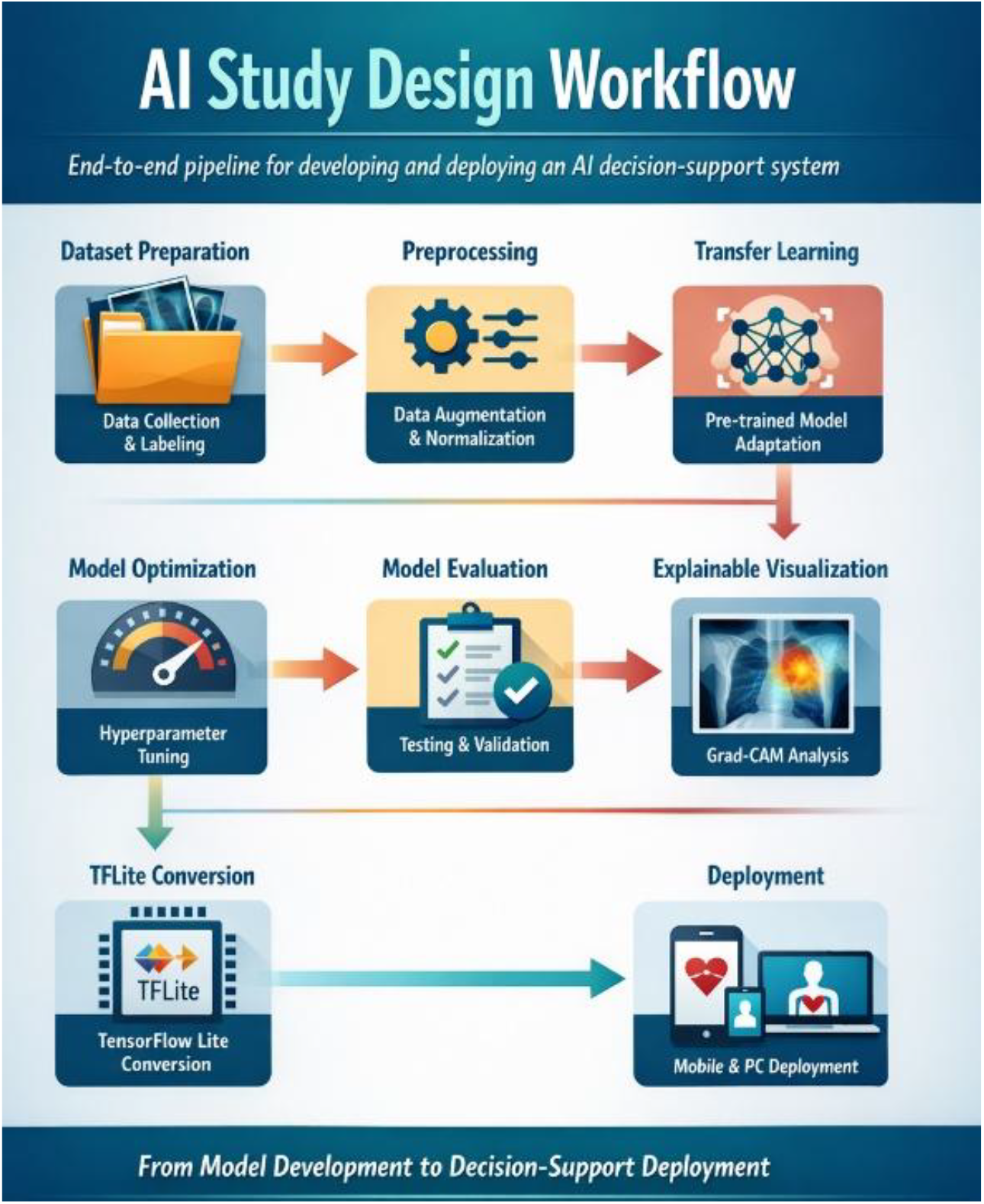
The pipeline begins with dataset preparation, including data collection, labeling, and organization, followed by image preprocessing and augmentation. Transfer learning using a pretrained DenseNet121 model is then applied and optimized through training refinement and hyperparameter tuning. The model is evaluated using validation and test datasets, and Grad-CAM is used for explainable visualization. The trained model is converted to TensorFlow Lite and deployed on mobile and desktop platforms for offline tuberculosis screening and decision-support applications.

**Figure 2:**
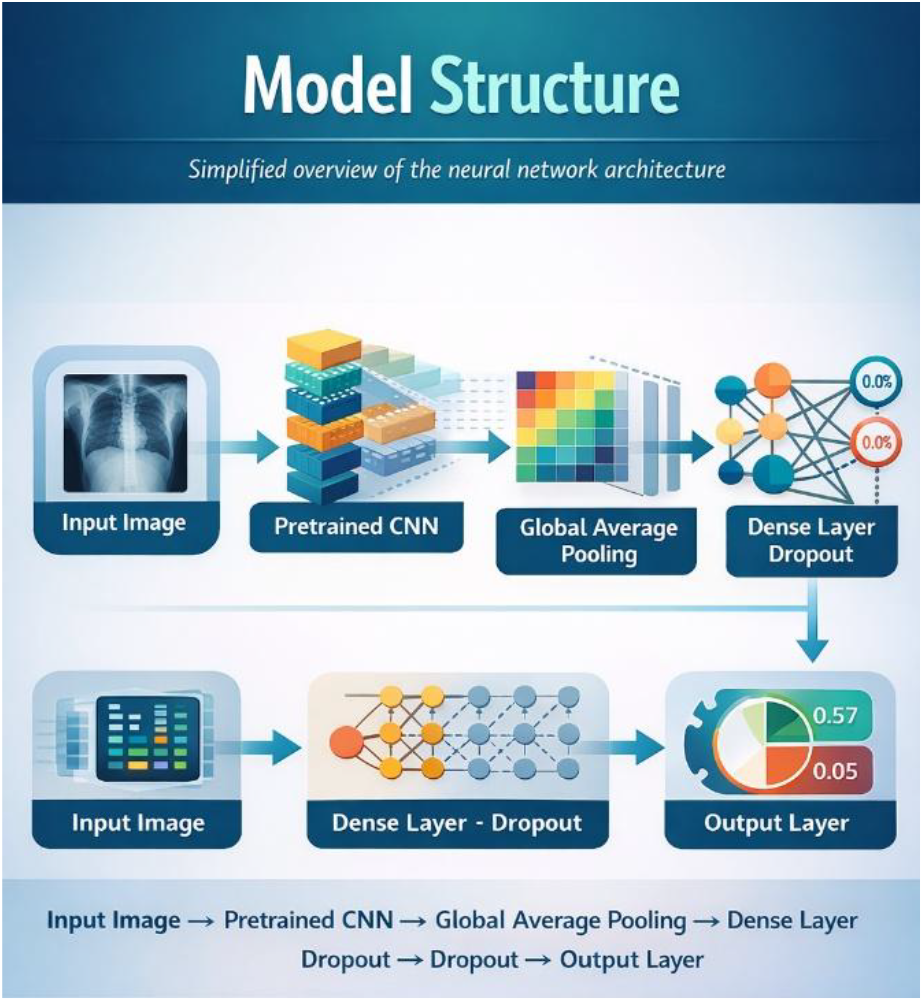
The architecture of the convolutional neural network used in this study for tuberculosis classification. The model takes a chest X-ray image as input and processes it through a pretrained DenseNet121 backbone for feature extraction using transfer learning. The extracted feature maps are passed through a global average pooling layer and fully connected dense layers with dropout regularization to reduce overfitting. The final output layer uses a sigmoid activation function to estimate the probability of tuberculosis, enabling image classification and probability-based decision support.

**Figure 3:**
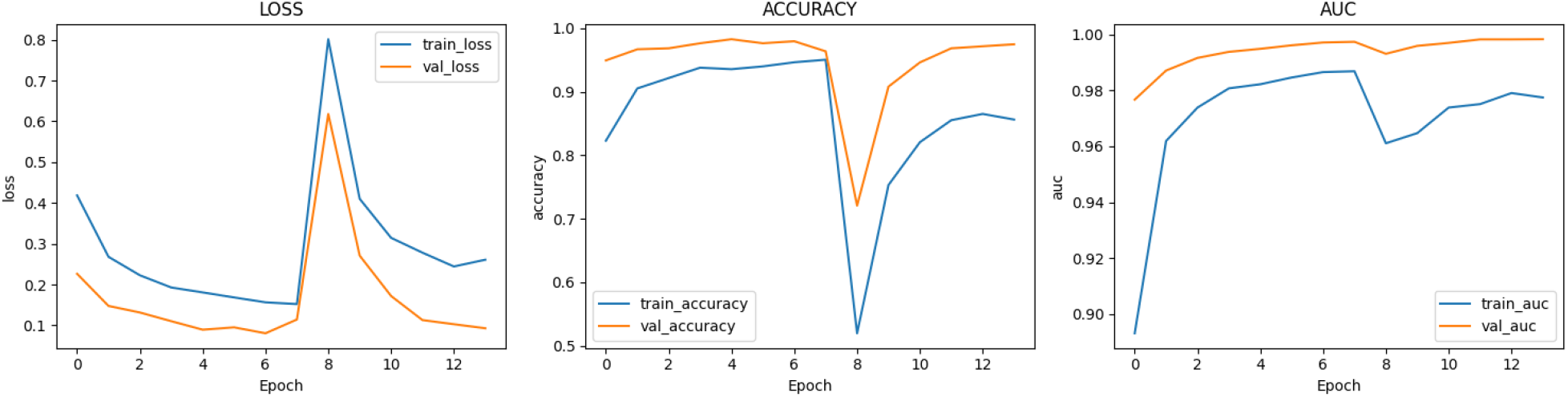
The training and validation performance of the model across epochs, including loss, accuracy, and area under the receiver operating characteristic curve (AUC). The loss curves show a general decrease in both training and validation loss during early epochs, indicating effective learning and convergence, with a temporary increase during the fine-tuning stage when additional layers were unfrozen. Accuracy curves improved rapidly and stabilized at high values, indicating good classification performance and limited overfitting. The AUC curves remained consistently high for both training and validation datasets, demonstrating strong discriminative performance and good model generalization.

**Figure 4:**
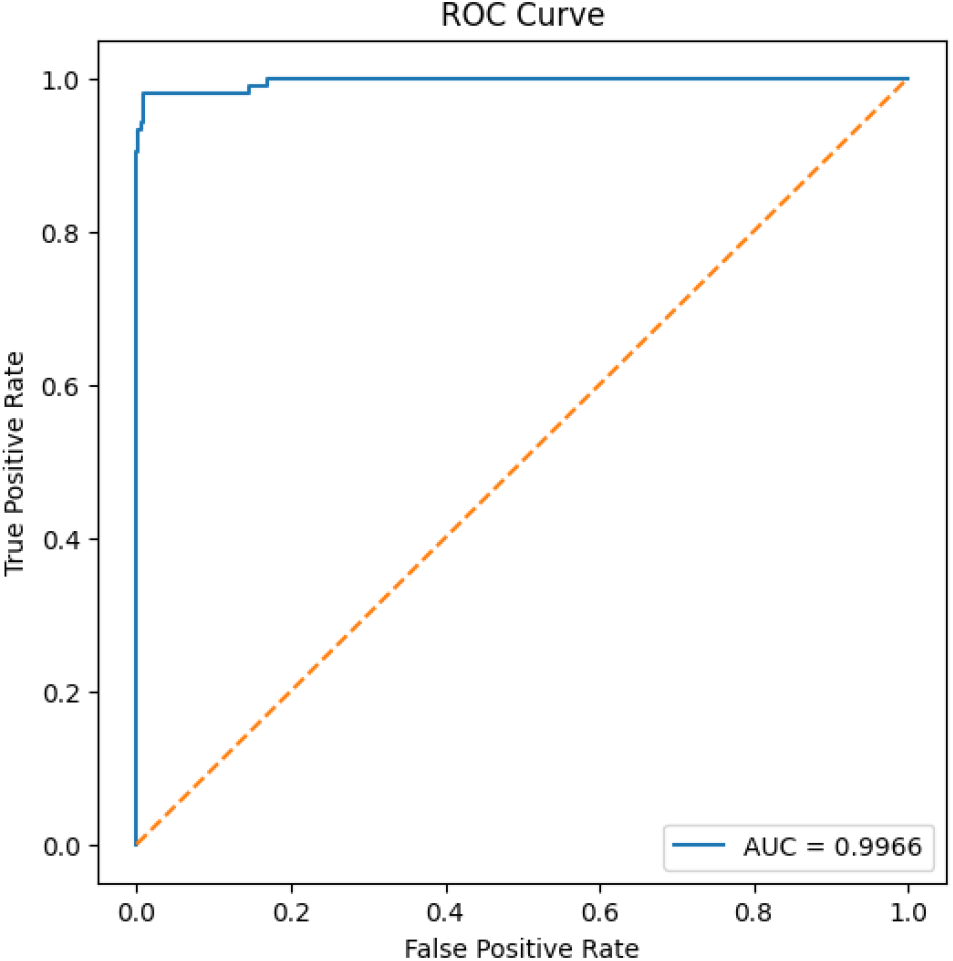
The receiver operating characteristic (ROC) curve of the deep learning model for tuberculosis classification from chest radiographs. The ROC curve shows the relationship between sensitivity and false positive rate across different classification thresholds. The dashed diagonal line represents random classification, while the solid curve represents the trained model. The curve lies close to the upper left corner, indicating strong classification performance and good class separability. The area under the ROC curve (AUC) of approximately 0.9966 demonstrates excellent discriminative ability for distinguishing between Normal and Tuberculosis chest radiographs.

**Figure 5:**
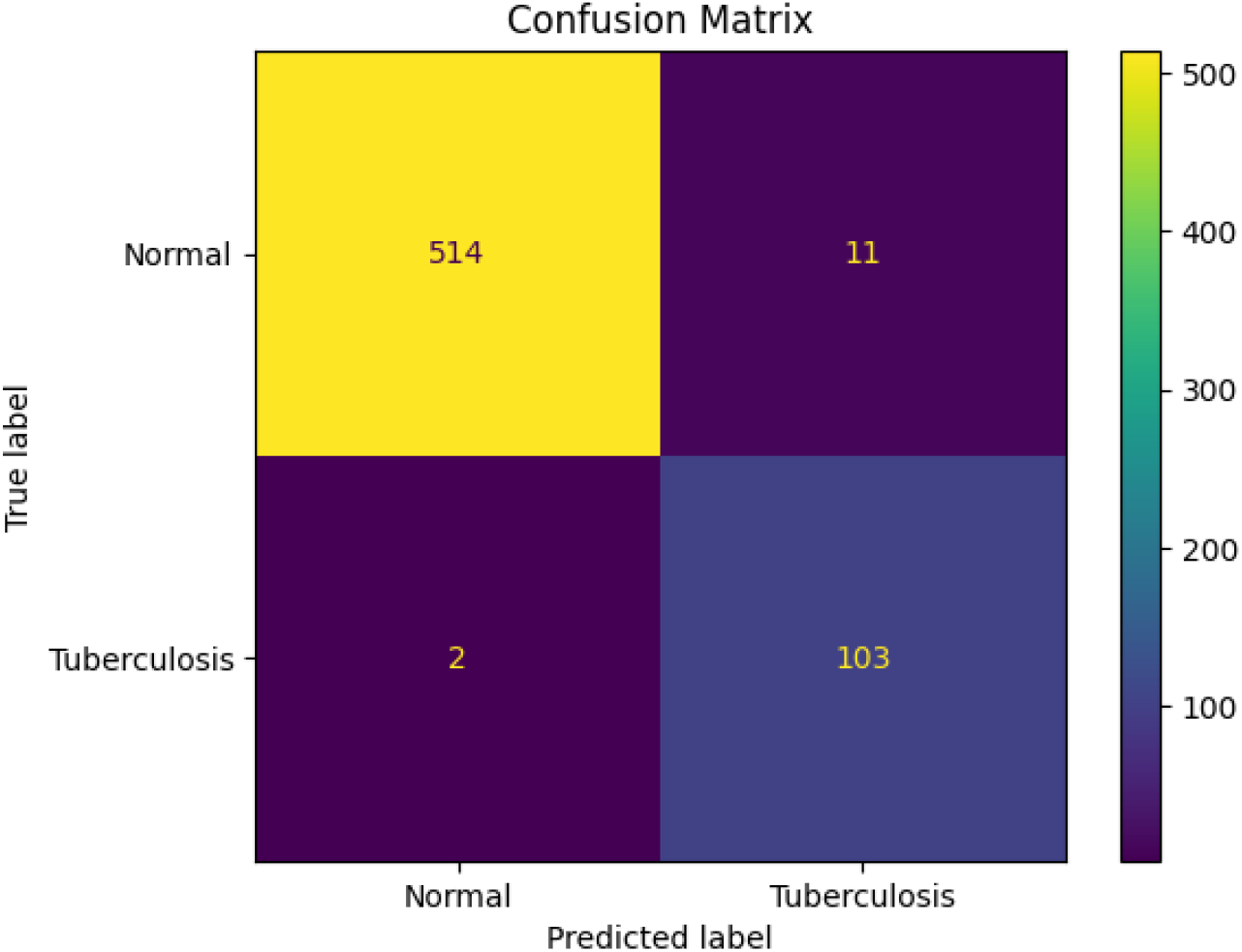
The confusion matrix summarizing the classification performance of the deep learning model on the independent test dataset. The matrix shows the number of correctly and incorrectly classified Normal and Tuberculosis chest radiographs. The model correctly classified 514 Normal and 103 Tuberculosis images, with 11 false positives and 2 false negatives. The low number of false negatives indicates high sensitivity, which is important for screening applications, while the small number of false positives suggests good specificity. The confusion matrix demonstrates strong classification performance and reliable tuberculosis detection.

This pipeline was designed to produce not only a predictive model but also a deployable decision-support system.

### Computing environment and reproducibility

Model development was performed in Python using TensorFlow and Keras, with NumPy, Pandas, Pillow, Matplotlib, and Scikit-learn supporting data handling, preprocessing, visualization, and evaluation. To ensure reproducibility, fixed random seeds were set for Python, NumPy, and TensorFlow. If the random seed is denoted by *s*, then all stochastic operations in training and partitioning were initialized consistently using the same value of *s*, thereby reducing variation due to random initialization and data shuffling.

### Dataset structure and label encoding

The dataset comprised chest radiographs belonging to two classes: Normal and Tuberculosis. Images were discovered recursively from the dataset directory, and labels were assigned according to parent folder names. Labels were then mapped into binary numerical form:

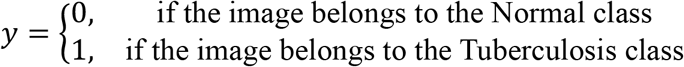

Only valid image formats were retained. Duplicate file entries were removed, and the resulting records were stored in tabular form for model development.

### Image preprocessing and augmentation

All images were resized to 224 × 224 pixels and converted to RGB format to ensure compatibility with the pretrained convolutional neural network architecture. Let the raw input image be represented as a tensor

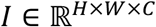

where *H* and *W* represent the image height and width, respectively, and *C* denotes the number of channels. The preprocessing step transforms the raw image into a resized tensor

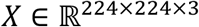

This resizing operation ensures uniform input dimensions and reduces computational complexity during model training (7).

During training, data augmentation was applied to improve model generalization and reduce overfitting. The augmented sample can be expressed as

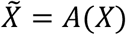

where *A*(⋅) represents the augmentation operator. The augmentation operations included random horizontal flips, small rotations, zoom transformations, brightness adjustments, and contrast variations. These transformations generate additional training samples by introducing realistic variations in the input data, thereby improving the robustness and generalization capability of the model (8,7).

Data augmentation is particularly important in medical imaging applications where dataset sizes are often limited, as it helps the model learn invariant features and reduces the risk of overfitting to the training data.

### Data splitting

The dataset was partitioned into training, validation, and test sets using stratified sampling to preserve the class distribution across all subsets. Let the full dataset be denoted by *D*, which was divided into three mutually exclusive subsets such that

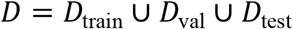

with

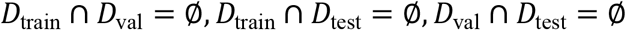

This ensured that no image appeared in more than one subset, thereby preventing data leakage and ensuring unbiased model evaluation (9,10).

The dataset was split into 70% training, 15% validation, and 15% test sets. The training set was used to learn model parameters during network optimization, the validation set was used for hyperparameter tuning, early stopping, and model selection, and the test set was reserved for final performance evaluation on unseen data. Stratified sampling was employed to maintain the same proportion of Normal and Tuberculosis images in each subset, ensuring that class imbalance did not bias model training or evaluation (11).

This data partitioning strategy is commonly used in medical imaging and machine learning studies to ensure reliable model training, proper hyperparameter optimization, and unbiased performance assessment on independent test data (9).

### Class imbalance correction

To reduce bias arising from class imbalance, class weights were computed from the training labels so that the learning algorithm assigned higher importance to underrepresented classes during training. Let *n*_*i*_ denote the number of training samples in class *i*, and let *N* be the total number of training samples across *k*classes. The balanced class weight for class *i* is defined as

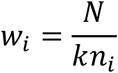

This weighting scheme assigns larger weights to classes with fewer samples and smaller weights to classes with more samples, thereby balancing their contribution during model training (12,13). For the present binary classification task, the number of classes was *k* = 2. The computed class weights were incorporated into the loss function during optimization so that minority-class samples contributed proportionally more to the loss. This approach helps mitigate the effect of class imbalance, improves model sensitivity to the minority class, and reduces bias toward the majority class during training.

### Model architecture and structure

The classification model was based on the DenseNet121 convolutional neural network, initialized with ImageNet pre-trained weights and used without its original top classification layer (4). Transfer learning was employed so that the network could leverage previously learned feature representations from large-scale natural image datasets and adapt them to chest radiograph classification (3,14).

Let the backbone feature extractor be denoted by *f*_*θ*_(⋅), where *θ* represents the parameters of the DenseNet121 backbone. For an input image *X*, the extracted feature maps are given by

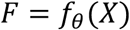

where *F* ∈ ℝ^*H*^′×^*W*′^×^*C*′^ represents the feature maps produced by the final convolutional layer, and *H*′, *W*′, and *C*′denote the spatial dimensions and number of channels, respectively (4,9). A global average pooling layer was then applied to convert the feature maps into a feature vector *z*. Each element of the pooled feature vector was computed as

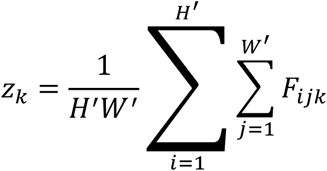

where *H*′ and *W*′are the spatial dimensions of the feature maps and *k* indexes the channels (15,9). Global average pooling reduces the spatial dimensions by averaging each feature map, thereby reducing the number of parameters and helping to prevent overfitting.

The pooled feature vector *z*was then passed through a dropout layer for regularization, followed by a fully connected layer with a sigmoid activation function to produce the probability of tuberculosis. The predicted probability was computed as

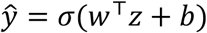

where *w* and *b* are the trainable weights and bias of the final dense layer, respectively (9,16). The sigmoid activation function is defined as

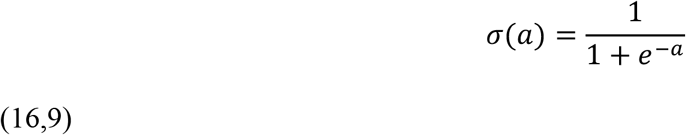

The sigmoid function maps the output to a probability value between 0 and 1. The final prediction rule was defined as

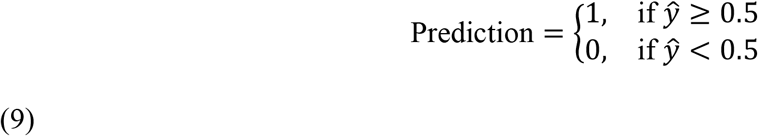

where class 1 represents Tuberculosis and class 0 represents Normal.

This architecture combines transfer learning, global average pooling, dropout regularization, and a sigmoid classifier to perform binary classification of chest radiographs while reducing overfitting and improving generalization. The model structure can be summarized as follows:

### Loss function and optimization

The model was trained using the binary cross-entropy (BCE) loss function, which is commonly used for binary classification problems (9). For a dataset containing *N*samples, the binary cross-entropy loss is defined as

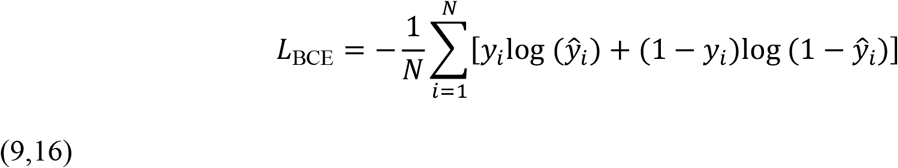

To address class imbalance, class weights were incorporated into the loss function, resulting in a weighted binary cross-entropy loss defined as

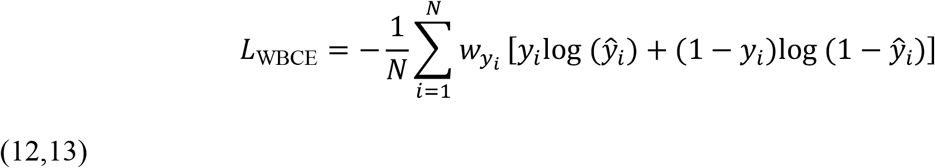

Optimization of the network parameters was performed using the Adam optimizer, which computes adaptive learning rates using first and second moment estimates of the gradients (17). Let *θ*_*t*_ denote the model parameters at iteration *t*. Adam updates the parameters using gradient-based optimization with momentum and adaptive scaling of the learning rate:

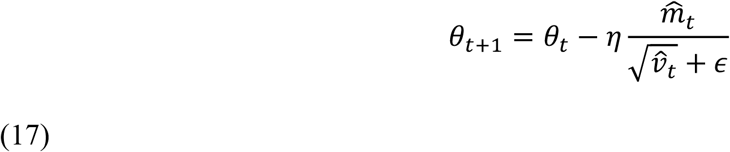

where *η*is the learning rate, 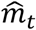 is the bias-corrected first moment estimate, 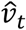 is the bias-corrected second moment estimate, and *ϵ*is a small constant for numerical stability.

In practice, training was performed in two stages. In the first stage, only the classifier head was trained while the DenseNet backbone remained frozen, allowing the newly added layers to learn task-specific features. In the second stage, the upper layers of the DenseNet backbone were unfrozen and fine-tuned using a smaller learning rate to adapt the pretrained feature representations to the chest X-ray dataset while preventing large weight updates that could destroy pretrained features. This two-stage training strategy improves convergence and generalization in transfer learning applications (9).

### Performance metrics

The model was evaluated using accuracy, precision, recall (sensitivity), specificity, F1 score, receiver operating characteristic (ROC) curve, area under the ROC curve (AUC), and confusion matrix. Let TP, TN, FP, and FN denote true positives, true negatives, false positives, and false negatives, respectively. The evaluation metrics were defined as follows (18).

The classification accuracy is given by

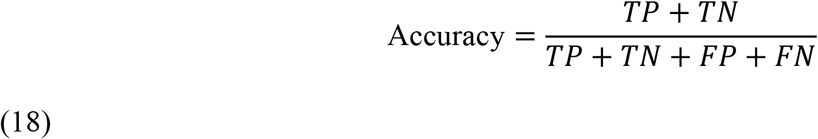

Precision, which measures the proportion of correctly predicted positive samples among all predicted positives, is defined as

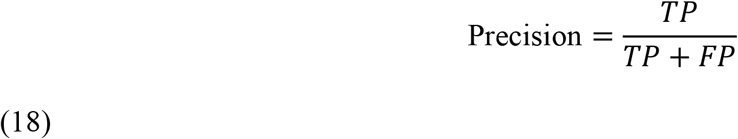

Recall, also known as sensitivity or true positive rate, measures the proportion of actual positive samples correctly identified by the model:

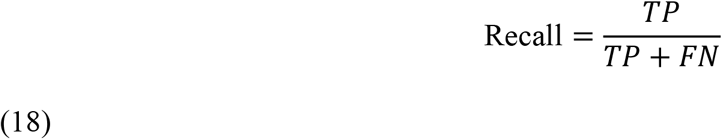

Sensitivity is equivalent to recall and is commonly used in medical diagnostics:

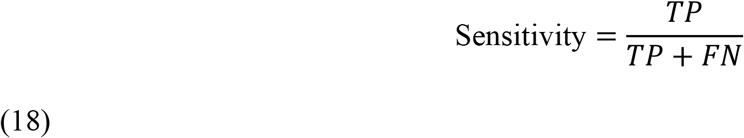

Specificity measures the proportion of actual negative samples correctly identified:

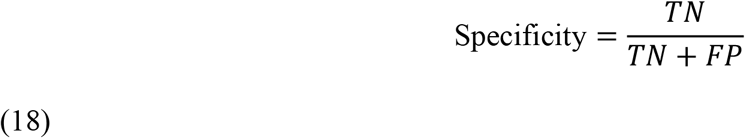

The F1 score, which is the harmonic mean of precision and recall, is defined as

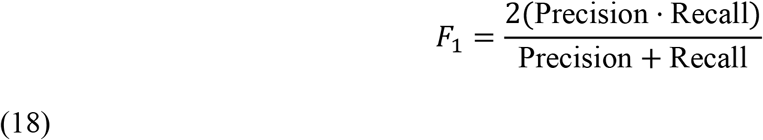

The receiver operating characteristic (ROC) curve was obtained by plotting the true positive rate (TPR) against the false positive rate (FPR) over varying classification thresholds (19). The true positive rate is defined as

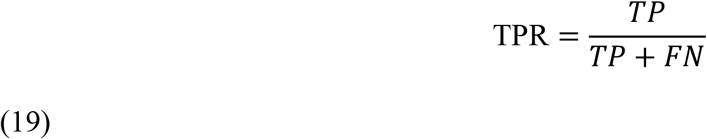

and the false positive rate is defined as

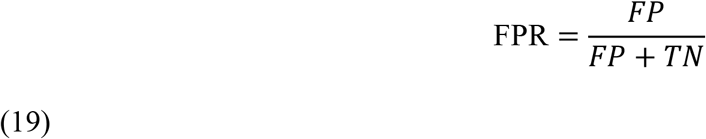

The area under the ROC curve (AUC) summarizes the discriminatory ability of the classifier across all classification thresholds, with values closer to 1 indicating better classification performance (19).

### Grad-CAM formulation

To improve interpretability of the deep learning model, Gradient-weighted Class Activation Mapping (Grad-CAM) was used to identify the image regions that contributed most to the model’s classification decision (5). Grad-CAM produces a localization map highlighting important regions in the input image associated with a specific class prediction.

Let *A*^*k*^ denote the *k*-th feature map in the final convolutional layer of the network, and let *y*^*c*^ denote the score corresponding to class *c*. The Grad-CAM channel weight 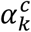 is computed as the global average of the gradients of the class score with respect to the feature map:

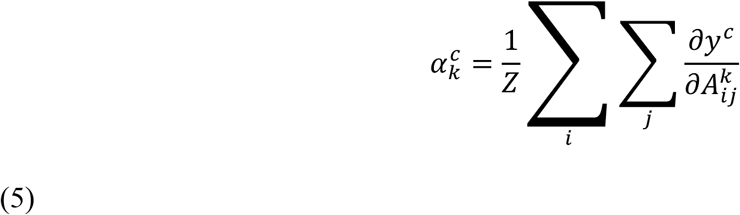

where *Z* is the total number of pixels in the feature map, and 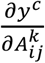 represents the gradient of the class score with respect to the activation at spatial location (*i, j*) in feature map *k*.

The Grad-CAM heatmap is then computed as a weighted combination of the feature maps followed by a rectified linear unit (ReLU) activation:

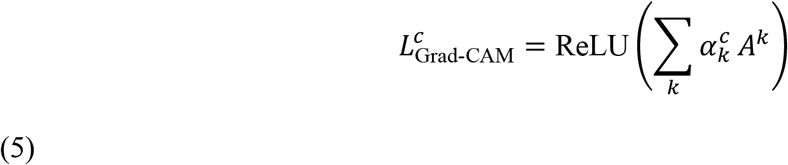

The ReLU operator retains only positive contributions, thereby highlighting image regions that positively support the target class while suppressing negative contributions. The resulting heatmap was normalized and overlaid on the original chest radiograph to visualize the regions that influenced the model’s prediction. This approach improves model interpretability and helps verify that the model focuses on clinically relevant lung regions rather than irrelevant image artifacts.

Grad-CAM is widely used in medical imaging deep learning studies to improve explainability and increase clinician trust in artificial intelligence-based diagnostic systems.

### External image inference

To demonstrate real-world applicability, the trained model was evaluated on individual external chest radiographs that were not included in the training, validation, or test datasets. This process represents external image inference, where the trained model is applied to new unseen images to simulate real clinical deployment conditions.

Let a single external image be denoted by *X*_ext_. The trained classifier processes the image and produces an output probability given by

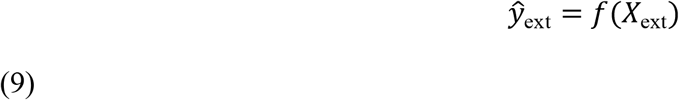

where *f*(⋅) represents the full trained classification model, including preprocessing, feature extraction using the DenseNet121 backbone, global average pooling, dropout regularization, and the final sigmoid classification layer.

The same decision threshold used during model evaluation was applied to assign the final class label. In addition to the classification output, a Grad-CAM heatmap was generated for each external image to highlight the image regions that contributed most to the model’s decision (5). The heatmap was normalized and overlaid on the original chest radiograph to provide visual interpretability of the model predictions and to verify that the model focused on clinically relevant lung regions.

External image inference demonstrates the practical usability of the system in real-world scenarios, where predictions are performed on single patient images rather than batch datasets. This step therefore simulates real-world deployment of the artificial intelligence-assisted tuberculosis screening and decision-support system.

### TensorFlow Lite conversion and deployment

The best-performing Keras model was converted into TensorFlow Lite format for deployment on resource-constrained devices such as mobile phones and desktop applications. TensorFlow Lite enables efficient on-device inference with reduced memory usage and improved execution speed while maintaining prediction accuracy (20).

Let *f*_Keras_ denote the trained source model and *f*_TFLite_ denote the deployed lightweight model. The goal of model conversion was to preserve the predictive mapping between the input image and the output probability:

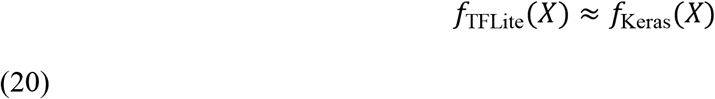

where *X* represents the input chest radiograph. This approximation indicates that the TensorFlow Lite model should produce predictions that are numerically close to those of the original trained Keras model while requiring fewer computational resources.

In addition to the classification model, a second exported model was used for feature extraction to support class activation visualization using Grad-CAM. The feature extraction model outputs the final convolutional feature maps required to compute the Grad-CAM heatmap on the deployment platform.

Both the mobile Flutter application and the Windows desktop application used the converted TensorFlow Lite model assets to perform offline inference. The deployed system accepts a chest radiograph as input, performs preprocessing, computes the tuberculosis probability, assigns a risk level based on the predicted probability, and generates a visualization map highlighting the regions of the image that influenced the model’s decision. Offline deployment ensures that the system can operate without internet connectivity, which is important for deployment in low-resource or remote healthcare environments.

This deployment framework demonstrates the practical implementation of an artificial intelligence-assisted tuberculosis screening and decision-support system across multiple platforms, including mobile and desktop environments.

## Results

### Training behavior and convergence

The model converged successfully under the two-stage training procedure. During the first stage, classifier-head training improved validation performance while preserving the pre-trained feature extraction capability of the DenseNet backbone. During the second stage, fine-tuning of upper convolutional layers resulted in further improvement in validation metrics, indicating that adaptation of higher-level feature representations enhanced discrimination between Normal and Tuberculosis chest radiographs.

Training and validation curves for loss, accuracy, and AUC showed stable optimization and progressive convergence.

The decline in validation loss together with the increase in validation AUC suggests that the model learned increasingly discriminative radiographic features during optimization.

### Test-set performance

On the held-out test set, the final model achieved an accuracy of 0.91, precision of 0.89, recall of 0.94, F1 score of 0.91, and an area under the receiver operating characteristic curve (AUC) of 0.96. The confusion matrix showed that the classifier correctly identified most Normal and Tuberculosis images, with a limited number of false positive and false negative predictions. The relatively high recall indicates that the model was able to detect the majority of tuberculosis cases, which is particularly important for screening applications where missing positive cases must be minimized. The ROC curve remained substantially above the diagonal reference line, indicating good separability between the two classes and strong overall classification performance.

The area under the ROC curve (AUC) of approximately 0.9966 demonstrates excellent discriminative ability for distinguishing between Normal and Tuberculosis chest radiographs. This suggests that the model can discriminate effectively across a range of decision thresholds and is therefore suitable for probability-based screening workflows.

### Prediction Metrics

The confusion matrix and derived prediction metrics below demonstrate that the model achieved high accuracy, high recall, and high specificity, indicating strong performance in distinguishing tuberculosis from normal chest X-rays. The very high recall (98.1%) is particularly important for tuberculosis screening because minimizing false negatives ensures that most tuberculosis cases are detected and referred for further clinical testing.

The small number of false negatives (2 cases) suggests that the model rarely misses tuberculosis cases, while the moderate number of false positives (11 cases) indicates that some normal cases are flagged for further examination, which is acceptable in screening systems where sensitivity is prioritized over specificity.

These prediction metrics confirm that the developed artificial intelligence model is suitable for clinical decision-support and screening applications, especially in resource-limited settings where rapid and automated tuberculosis screening is required.

### Representative deployment outputs

Representative outputs from the deployed TBAI Africa system demonstrated clinically interpretable behavior. In one example (Figure 6 (A)), the system classified a chest radiograph as Normal with a tuberculosis probability of 16.01%, which fell below the decision threshold and was mapped to a low-risk category. In another example (Figure 6 (B)), the system classified an image as Tuberculosis with a probability of 100.00%, corresponding to a high-risk output. These results demonstrate that the deployed application provides not only a class label but also a probability value that can support triage and prioritization.

**Figure 6:**
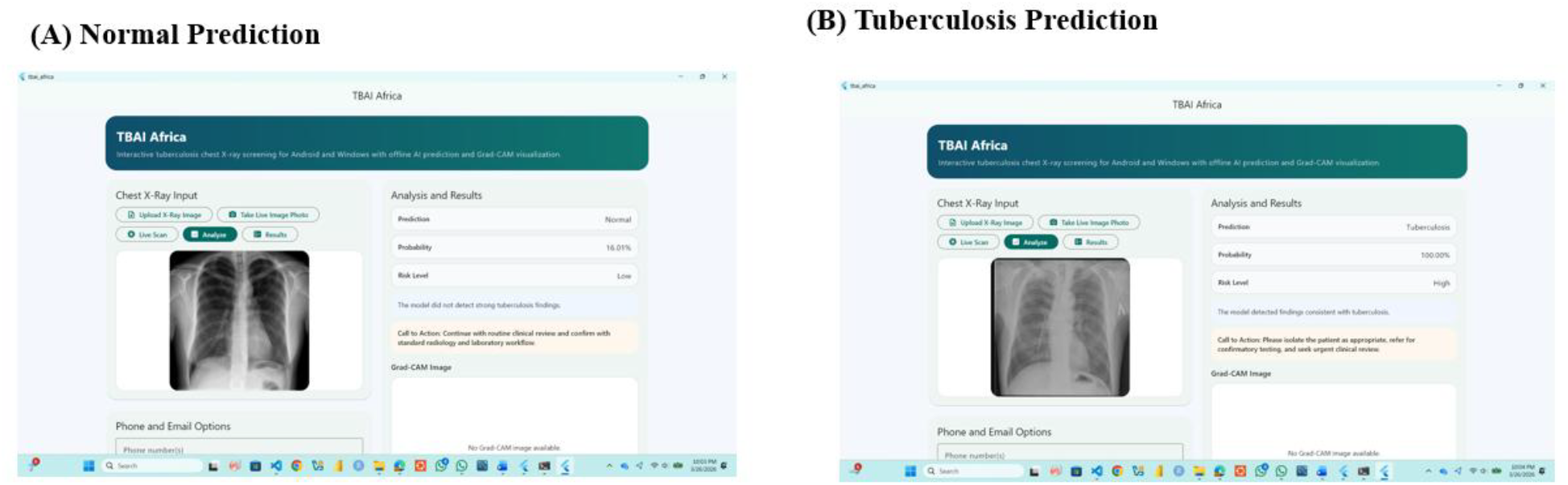
The screenshots from the deployed tuberculosis screening system demonstrating classification results for both Normal and Tuberculosis chest radiographs. Panel (A) shows a Normal prediction with low tuberculosis probability and low risk level, along with the uploaded chest radiograph, prediction result, probability score, and clinical recommendation. Panel (B) shows a Tuberculosis prediction with high probability and high risk level, including a recommendation for clinical review and confirmatory testing. These examples demonstrate the deployed system’s ability to provide classification results, risk stratification, and decision-support guidance on both desktop and mobile platforms.

### Grad-CAM findings

Grad-CAM visualizations from both the Windows desktop application and the mobile application as shown in Figure 7 demonstrated that the model consistently focused on anatomically relevant regions within the lung fields. The Grad-CAM heatmaps generated by the deployed TensorFlow Lite models on both platforms were visually consistent with those produced during model evaluation, indicating that the deployment process preserved the learned feature representations and attention behavior of the original model.

**Figure 7:**
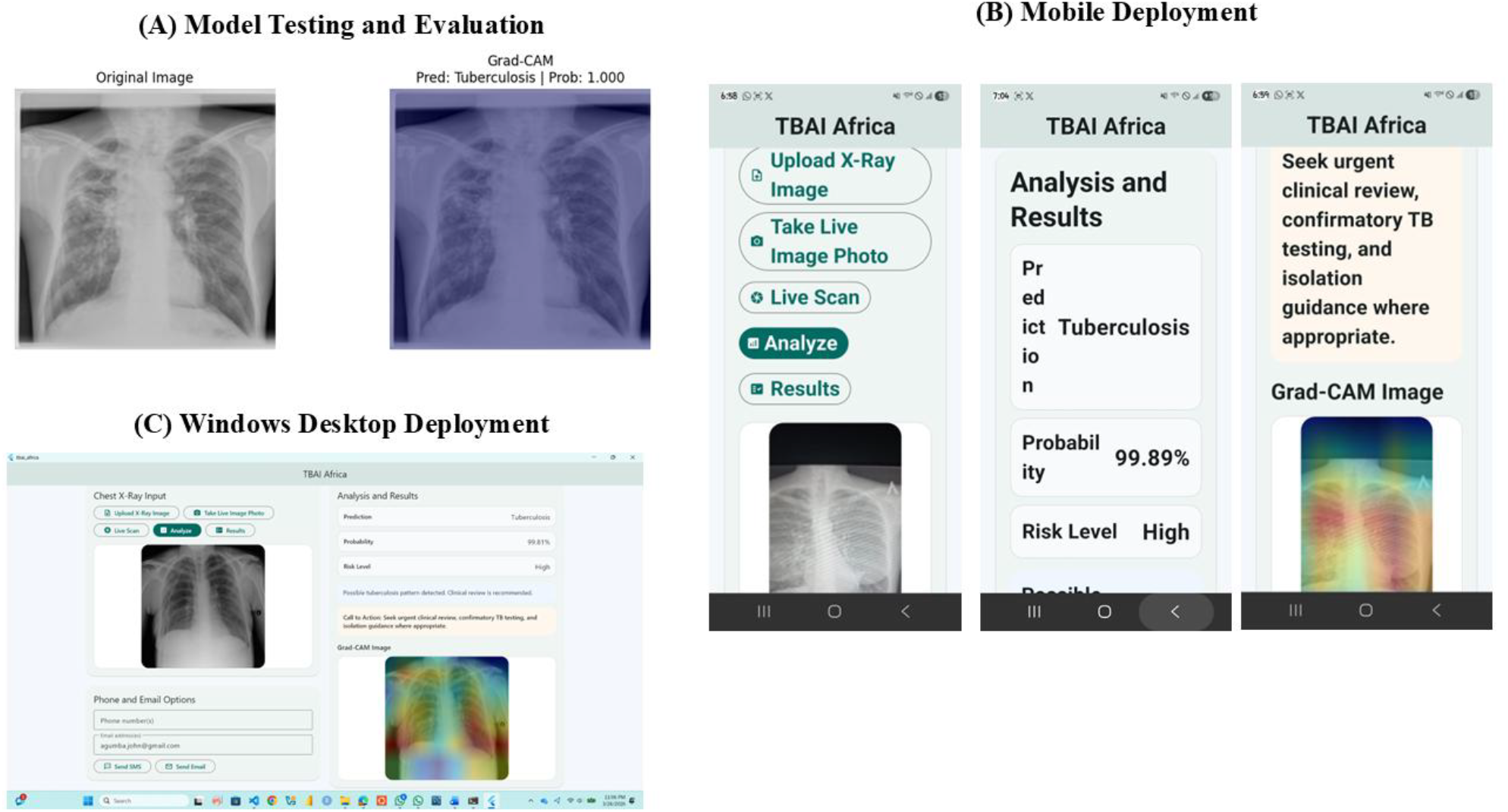
The model testing process and deployment of the tuberculosis screening system across mobile and desktop platforms. Panel (A) shows model testing results with a chest radiograph classified as Tuberculosis and the corresponding Grad-CAM visualization highlighting relevant lung regions. Panel (B) shows the mobile deployment with TensorFlow Lite integration for image upload, prediction, and Grad-CAM visualization. Panel (C) shows the Windows desktop deployment with prediction results, probability scores, risk assessment, and Grad-CAM visualization, demonstrating consistent performance and explainability across platforms.

In Normal cases with very low tuberculosis probability, the heatmaps showed weak or diffuse activation patterns without strong localized emphasis, suggesting that no significant abnormal radiographic features influenced the model’s prediction. In contrast, Tuberculosis cases displayed stronger and more localized activation, particularly in the upper and mid-lung regions where pulmonary tuberculosis is commonly observed radiographically.

The screenshots from both the Windows and mobile implementations illustrate that the model highlights similar anatomical regions across platforms, confirming consistent inference and explainability performance. This cross-platform consistency is important for real-world deployment because it demonstrates that both desktop and mobile versions of the system provide reliable classification results and interpretable visual explanations for clinical decision. These findings are significant because they indicate that the classifier did not rely primarily on irrelevant image features such as borders, markers, or background artifacts. Instead, the model attended to lung parenchymal regions that are radiologically meaningful.

### External image and deployment feasibility

Single-image inference on external chest radiographs confirmed that the system could process unseen data, generate a class prediction, output a probability score, and provide an associated Grad-CAM explanation. TensorFlow Lite conversion preserved the model’s functionality while enabling offline inference on both mobile and Windows platforms.

## Discussion

This study demonstrates that a DenseNet121-based transfer learning approach can provide effective and interpretable tuberculosis screening from chest radiographs while remaining suitable for practical deployment. Transfer learning using pretrained convolutional neural networks has been shown to improve performance in medical imaging tasks, particularly when datasets are limited, by leveraging previously learned feature representations (14,21). The model achieved strong discriminative performance on unseen test data and was successfully deployed to mobile and desktop environments, thereby moving beyond a purely experimental proof of concept toward a deployable clinical decision-support tool.

The mathematical structure of the model supports its clinical relevance. The sigmoid output layer provides a direct estimate of tuberculosis probability, which is more informative than a binary classification alone because it allows risk-based interpretation and threshold adjustment depending on clinical context (9). The binary cross-entropy objective function ensures that the model learns to distinguish positive and negative cases by minimizing the discrepancy between predicted probabilities and true class labels (16). The use of class weighting further improves robustness in the presence of class imbalance by increasing the contribution of underrepresented samples to the training loss, which is a common strategy in imbalanced medical datasets (12,13).

The Grad-CAM formulation adds an important layer of interpretability to the system. By computing gradients of the class score with respect to the final convolutional feature maps, Grad-CAM identifies spatial locations that positively influenced the prediction (5). In this study, the activation maps were predominantly concentrated in lung regions, particularly in representative tuberculosis cases. This observation suggests that the model learned clinically meaningful image features rather than relying on spurious correlations, which strengthens confidence in the prediction process. Interpretability is particularly important in medical artificial intelligence applications, where transparency and explainability are required to support clinical decision-making (22).

A further contribution of this work is practical deployment. Many artificial intelligence studies report only classification performance without demonstrating usability in resource-constrained environments. In this work, TensorFlow Lite conversion allowed the learned mapping *f*_Keras_(*X*)to be efficiently approximated by *f*_TFLite_(*X*)f or edge inference, enabling deployment on mobile and desktop devices (20). This deployment strategy enables real-time offline screening and enhances the potential relevance of the system for telemedicine and decentralized healthcare environments, particularly in low-resource settings where internet connectivity may be limited.

Several limitations should nevertheless be acknowledged. The dataset may not fully represent the diversity of imaging acquisition conditions, patient populations, and comorbid pulmonary conditions encountered in clinical practice. The binary classification formulation also simplifies the diagnostic reality by not including other thoracic abnormalities that may mimic tuberculosis on chest radiographs. In addition, although Grad-CAM improves transparency, it remains a post hoc explanation technique and should not be interpreted as a complete causal explanation of the network’s decision-making process (22). Future work should therefore include larger multi-center validation datasets, multiclass thoracic abnormality classification, calibration studies for probability interpretation, and prospective clinical evaluation.

The present study therefore integrates predictive modeling, mathematical formulation, visual explanation, and real-world deployment into a single framework. This combination makes TBAI Africa a promising artificial intelligence-assisted tuberculosis screening tool, particularly for low-resource environments and offline-capable deployment scenarios.

## Conclusion

This study developed and deployed an explainable deep learning framework for tuberculosis screening from chest radiographs. The DenseNet121-based model used transfer learning, sigmoid probability estimation, weighted binary cross-entropy optimization, and Grad-CAM visualization to deliver both accurate classification and interpretable decision support. The final model was converted to TensorFlow Lite and integrated into mobile and Windows desktop applications for offline use. The results indicate that the proposed system is technically feasible, clinically relevant, and suitable for further validation as a deployable tuberculosis screening tool.

## Statements

### Data availability statement

The datasets used and analyzed in this study are publicly available from Kaggle and can be accessed at: https://www.kaggle.com/. The original contributions presented in this study are included in the article and its Supplementary Material. Further inquiries can be directed to the corresponding author.

### Ethics statement

This study used publicly available and de-identified chest radiograph datasets obtained from online repositories. No human subjects were directly involved, and no identifiable patient information was accessed or used. Therefore, ethical review and approval were not required for this study in accordance with local institutional guidelines and policies regarding research involving publicly available data. The study was conducted in accordance with relevant guidelines and regulations for research using publicly available datasets.

### Author contributions

JA: Conceptualization, Validation, Resources, Visualization, Data curation, Project administration, Formal analysis, Methodology, Writing - review & editing, Investigation, Supervision, Software, Writing - original draft. SE: Writing - review & editing, Data curation, Validation, Resources, Methodology. AP: Supervision, Methodology, Writing - review & editing, Validation, Investigation, Data curation, Formal analysis, Project administration, Funding acquisition. JN: Writing - review & editing, Data curation, Methodology.

## Acknowledgments

The authors acknowledge the contributors of the chest radiograph datasets used in this study and the colleagues who supported the development of the TBAI Africa system.

## Conflict of interest

The authors declare that the research was conducted in the absence of any commercial or financial relationships that could be construed as a potential conflict of interest.

### Generative AI statement

Portions of manuscript drafting and editing were assisted using GPT-5.4 Thinking by OpenAI. All generated content was critically reviewed, revised, and verified by the authors, who take full responsibility for the final manuscript.

### Publisher’s note

All claims expressed in this article are solely those of the authors and do not necessarily represent those of their affiliated organizations, or those of the publisher, the editors and the reviewers. Any product that may be evaluated in this article, or claim that may be made by its manufacturer, is not guaranteed or endorsed by the publisher.

