## Supplementary Material for "A Deployable Explainable Deep Learning System for Tuberculosis Detection from Chest X-Rays in Resource-Constrained High-Burden Settings"

### Supplementary Material S1. Additional methodological details

#### S1.1 Dataset organization and label harmonization

The dataset used in this study consisted of chest radiograph images organized into folder structures representing diagnostic categories. Because source datasets may use slightly different folder names for the tuberculosis class, a label harmonization procedure was implemented before model training. Folder names were mapped to the unified class label **Tuberculosis**, while the folder name *normal* was mapped to the unified class label **Normal**. This harmonization step ensured consistency in binary class labeling across all data sources.

#### S1.2 Image discovery and record creation

Image files were discovered recursively from the dataset root directory. Only valid image extensions were retained, including PNG, JPG, JPEG, BMP, and WEBP formats. For each detected image, the file path and inferred class label were stored in a structured tabular record. Duplicate file paths were removed prior to training. A validation check was then applied to confirm that both target classes were represented in the final dataset.

#### S1.3 Image preprocessing

All images were converted to RGB format and resized to $224 \times 224$ pixels before being passed to the deep learning model. Pixel arrays were stored as floating-point tensors. This preprocessing step standardized the image input dimensions and ensured compatibility with the DenseNet121 architecture.

#### S1.4 Dataset partitioning

The curated dataset was divided into training, validation, and test subsets using stratified sampling to preserve class balance. The validation set represented 15% of the data and the test set represented 15%, leaving 70% of the data for model training. Stratified sampling was used to reduce the risk of class distribution shifts between subsets.

#### S1.5 Data augmentation

To improve generalization and reduce overfitting, augmentation was applied to the training data only. The augmentation pipeline included horizontal flipping, small rotations, zooming, random brightness adjustment, and random contrast variation. These transformations simulated modest radiographic variability while preserving the diagnostic content of the image.

#### S1.6 Class imbalance correction

To account for potential class imbalance, class weights were computed from the training set and passed to the optimization process. This ensured that the minority class contributed proportionally more to the loss function during training, thereby reducing bias toward the majority class.

#### S1.7 Model training procedure

The model training process consisted of two stages. In the first stage, the DenseNet121 backbone remained frozen and only the final classification head was trained. In the second stage, the upper portion of the convolutional backbone was unfrozen and fine-tuned using a smaller learning rate. This two-stage training strategy allowed the classifier to first adapt the output layer to the tuberculosis task and then refine higher-level visual features relevant to chest radiography.

#### S1.8 Callbacks and model selection

The following training callbacks were used:

- **Early stopping**, to terminate training when validation AUC no longer improved
- **Reduce learning rate on plateau**, to lower the learning rate when validation loss stagnated
- **Model checkpointing**, to save the best model according to validation AUC

The best saved checkpoint was used for all final evaluations and deployment export.

### Supplementary Material S2. Mathematical formulation

#### S2.1 Binary class encoding

The target variable was defined as

$$y = \begin{cases} 0, & \text{Normal} \\ 1, & \text{Tuberculosis} \end{cases}$$

#### S2.2 Sigmoid probability output

For an input image $X$, the classifier produced a tuberculosis probability

$$\hat{y} = \sigma(w^{\top} z + b),$$

where $z$ is the pooled feature vector, $w$is the dense-layer weight vector, $b$is the bias term, and $\sigma(\cdot)$ is the sigmoid function

$$\sigma(a) = \frac{1}{1 + e^{-a}}.$$

#### S2.3 Decision threshold

The final class prediction was assigned using a threshold of 0.5:

$$\text{Prediction} = \begin{cases} \text{Tuberculosis}, & \hat{y} \geq 0.5 \\ \text{Normal}, & \hat{y} < 0.5 \end{cases}$$

#### S2.4 Binary cross-entropy loss

Model optimization was based on the binary cross-entropy loss

$$\mathcal{L}_{\text{BCE}} = -\frac{1}{N}\sum_{i=1}^{N}[y_i \log(\hat{y}_i) + (1 - y_i)\log(1 - \hat{y}_i)]\,.$$

When class weights were used, the weighted form became

$$\mathcal{L}_{\text{WBCE}} = -\frac{1}{N}\sum_{i=1}^{N} w_{y_i}\,[y_i \log(\hat{y}_i) + (1 - y_i)\log(1 - \hat{y}_i)].$$

#### S2.5 Performance metrics

The performance metrics were defined as follows:

$$\text{Accuracy} = \frac{TP + TN}{TP + TN + FP + FN}$$
$$\text{Precision} = \frac{TP}{TP + FP}$$
$$\text{Recall} = \frac{TP}{TP + FN}$$
$$F_1 = 2 \cdot \frac{\text{Precision} \cdot \text{Recall}}{\text{Precision} + \text{Recall}}$$

#### S2.6 Grad-CAM formulation

For class $c$, the Grad-CAM weights were computed from the gradients of the class score $y^c$ with respect to the final convolutional feature maps $A^k$:

$$\alpha_k^c = \frac{1}{Z}\sum_i \sum_j \frac{\partial y^c}{\partial A_{ij}^k}$$

The class activation map was then given by

$$L^c_{\text{Grad-CAM}} = \text{ReLU}\left(\sum_k \alpha_k^c A^k\right)$$

where ReLU retains positive contributions associated with the target class.

**Supplementary Material S3. Model hyperparameters**

**Table S1. Training and deployment hyperparameters**

| **Parameter** | **Value** |
|---|---|
| Input image size | $224 \times 224$ |
| Batch size | 16 |
| Initial training epochs | 8 |
| Fine-tuning epochs | 6 |
| Initial learning rate | $1 \times 10^{-3}$ |
| Fine-tuning learning rate | $1 \times 10^{-5}$ |
| Validation split | 15% |
| Test split | 15% |
| Dropout rate | 0.3 |
| Backbone model | DenseNet121 |
| Output activation | Sigmoid |
| Classification threshold | 0.5 |

**Supplementary Material S4. Additional training details**

**S4.1 Software libraries**

The model development environment included:

- Python

- TensorFlow / Keras
- NumPy
- Pandas
- Matplotlib
- Pillow
- Scikit-learn

#### S4.2 Reproducibility settings

To improve reproducibility, the random seed was fixed across Python, NumPy, and TensorFlow. This reduced run-to-run variation in dataset splitting, shuffling, and model initialization.

#### S4.3 Output artifacts

The training and export process generated the following deployment artifacts:

- Trained Keras model file
- Label text file
- Main TensorFlow Lite classification model
- TensorFlow Lite feature extraction model for Grad-CAM support
- Metadata JSON file containing model input shape, output shape, feature output shape, and class labels

### Supplementary Material S5. TensorFlow Lite deployment details

#### S5.1 Mobile deployment

The trained model was converted from Keras format to TensorFlow Lite format for integration into a Flutter mobile application. The mobile application was designed to:

- load an existing chest radiograph from device storage or capture an image
- run offline inference on the device
- output the predicted class and probability score
- assign a risk category
- display Grad-CAM visual explanations

#### S5.2 Windows deployment

A Windows desktop interface was also developed using the exported model artifacts. The desktop application enabled:

- loading of chest radiograph images
- local prediction without internet connectivity
- display of class probabilities
- rendering of Grad-CAM overlays for interpretability

#### S5.3 Offline inference

Both the mobile and desktop versions were designed for offline inference, making the system appropriate for low-resource settings and environments where cloud access may be limited or unavailable.

### Supplementary Material S6. Code and implementation note

The codebase for model training, evaluation, Grad-CAM generation, TensorFlow Lite export, and deployment integration was developed as part of the TBAI Africa framework. The implementation included:

- automated dataset scanning and label assignment
- TensorFlow data pipelines for image loading and augmentation
- DenseNet121-based transfer learning
- two-stage optimization with fine-tuning
- Grad-CAM heatmap generation
- TensorFlow Lite export for edge deployment
- integration with mobile and desktop user interfaces

If the authors elect to provide code publicly, the repository link and version identifier should be inserted here.

**Code availability:** Available from the corresponding author upon reasonable request.

**Supplementary Material S7. AI use disclosure**

Portions of manuscript drafting and language refinement were assisted using GPT-5.4 Thinking by OpenAI. All text, scientific content, references, equations, and interpretations were critically reviewed, corrected where necessary, and approved by the authors, who take full responsibility for the final content of the manuscript and supplementary material.
